# Advancing Clinical Implementation of Cardiovascular Polygenic Risk Scores Through Patient-Level Robustness Assessment

**DOI:** 10.64898/2026.06.10.26355357

**Authors:** Roxane de La Harpe, Julien Vaucher, Zoltán Kutalik, Jacques Fellay, Christian W. Thorball

## Abstract

**Background and Aims:** Polygenic risk scores (PRSs) for atherosclerotic cardiovascular disease (ASCVD) can perform equivalently at the population level yet disagree for individual patients. We examined whether such intra-individual variability reflects genuinely complementary risk information or mainly statistical and methodological uncertainty, and whether it affects clinical classification once PRSs are integrated into SCORE2-OP.

**Methods:** In 4,137 ASCVD-free participants of the CoLaus|PsyCoLaus cohort (478 incident events over a median 14.4 years), we identified 16 ASCVD-PRSs with practically equivalent population-level performance using Bayesian equivalence testing. We quantified intra-individual variability (standard deviation, coefficient of variation, intraclass correlation, Cohen’s kappa, extreme discordance), tested whether discordance exceeded chance, decomposed scores into shared and unique genetic components, and assessed variability after integration into SCORE2-OP, benchmarked against perturbation of systolic blood pressure.

**Results:** For a typical individual, risk estimates varied by ±18 percentile points across PRSs. Discordance matched chance expectations under a shared-signal model, with no distinct phenotypic profile among discordant individuals, and predictive power resided overwhelmingly in the shared genetic component. Variability tracked PRS size and weighting rather than distinct variants. After integration into SCORE2-OP, 75.6% of participants were placed in different categories by at least one model and 54.6% as both low and high risk; instability was concentrated near guideline thresholds and far exceeded that from blood-pressure measurement error.

**Conclusions:** Equivalent population-level performance is not sufficient to treat PRSs as interchangeable at the individual level, and methodological standardisation and pragmatic clinical trials remain necessary to determine whether PRS integration improves long-term cardiovascular outcomes.

## Introduction

Polygenic risk scores (PRSs) for atherosclerotic cardiovascular disease (ASCVD) are gaining prominence as a valuable tool for risk prediction.^1,2^ They condense complex genetic information into a single metric. They represent lifelong probabilistic estimates of genetic risk based on population-level DNA associations,^3^ and favourable lifestyle choices can meaningfully mitigate the risk they convey.^4–6^ PRSs are now commercially available,^7^ several trials aim to validate their clinical utility,^8,9^ and medical societies have begun to issue statements on their use.^1,2^ More than 15 ASCVD-PRSs have been developed and validated, showing incremental value over traditional risk scores.^2^ In a Swiss population-based sample, we previously evaluated three ASCVD-PRSs. They showed similar predictive performance and added value in refining stratification, particularly at intermediate clinical risk.^10^ However, recent work has highlighted substantial individual-level variability in risk estimates, even among PRSs performing equivalently at the population level.^11–13^ One study of 46 CHD-PRSs found that 20% of participants had at least one PRS placing them in both the top and bottom 5th percentiles.^13^ Understanding this variability is essential for robust clinical implementation and for refining PRS selection.

In this study, we examined whether ASCVD-PRSs that perform similarly at the population level give consistent estimates for the same individual. The key question was whether intra-individual differences reflect meaningful additional risk information, such as distinct genetic pathways, or arise mainly from statistical and methodological uncertainty. We also assessed whether this variability affects clinical classification once PRSs are integrated into a guideline risk tool.

## Methods

### Study population

The CoLaus|PsyCoLaus study is a population-based study of the epidemiology and genetic determinants of cardiovascular and psychiatric disease in Lausanne, Switzerland.^14^ 6,733 individuals from the adult population of Lausanne were randomly recruited between 2003 and 2006 (age range 35–75 years, 54% women). They underwent clinical assessment and provided fasting blood samples, with longitudinal follow-up every five years to 2022 with ASCVD independently adjudicated by experts.^15^ Ethics committee of Canton Vaud gave ethical approval for this work (PB_2018-00038, ref. 239/09).

### Selection of participants

We included European-ancestry participants with quality-controlled genetic data and no ASCVD at baseline, excluding those lost to follow-up. For the fourth objective, participants with incomplete SCORE2/-OP inputs were also excluded.

### PRSs selection

PRSs were sourced from the Polygenic Score Catalog (February 2025).^16^ Of 70 ASCVD-PRSs initially considered, we excluded those not validated in European-ancestry samples or without added value beyond clinical scores in an independent cohort. PRSs from 2024/2025 were exempt, yielding 17 PRSs (*Supplementary Table 1, construction details in Supplementary Table 2 and Supplementary Methods*). We first retained only PRSs with equivalent population-level performance, so that individual-level differences would reflect differences in risk distribution rather than disparities in overall predictive accuracy. As an initial step, the 17 normalised PRSs were tested for association with incident ASCVD using Cox models adjusted for age, sex, and 10 principal components; those not nominally significant (p > 0.05) were excluded. Predictive performance (Brier score, AUROC) was then compared using the Bayesian approach of Abramowitz et al.^13^ Pairwise differences relative to a reference PRS were evaluated against a region of practical equivalence (ROPE ±0.02). PRSs with ≥95% of posterior samples within the ROPE were deemed equivalent (full procedure in Supplementary Methods).

### Intra-individual variability across ASCVD-PRSs with similar population-level performance

We used metrics replicating a recent study.^13^ We first used descriptive statistics to summarize how genetic risk estimates varied across individuals for similarly performing PRSs. For each individual we computed a mean risk percentile across PRSs. We then quantified within-person dispersion using the standard deviation (SD) and coefficient of variation (CV), with bootstrapped 95% CIs. We computed the intraclass correlation coefficient (ICC) for consistency across PRSs, and Light’s kappa for agreement in classifying individuals as high- (>90th/95th) or low-risk (<10th/5th). We also measured the proportion of individuals with PRSs in both extremes simultaneously. Among individuals with ≥3 extreme PRSs, we identified scores falling in the opposite extreme to the majority (full procedure in Supplementary Methods).

### Exploring sources of intra-individual variability across ASCVD-PRSs with equivalent population-level performance

PRSs for the same outcome often share only a small fraction of their SNPs. Two interpretations of intra-individual variability are therefore possible, and each has different implications for clinical implementation. Under the first interpretation, the distinct SNPs in each PRS genuinely capture complementary risk information, therefore the intra-individual variability between scores is itself informative. Under the alternative interpretation, the distinct SNPs represent different statistical attempts to capture the same underlying biological signal, and the variability reflects noise rather than information.

If this second interpretation holds, the noise can originate at two distinct levels. The first is the limited overlap in SNP composition across PRSs: when two PRSs use different SNPs to tag the same underlying genetic signal, those SNPs are not perfectly interchangeable, and these measurement differences accumulate across thousands of SNPs to produce discordant individual-level scores. The second concerns the SNP weights themselves, which can differ between PRSs for two reasons: (i) during the estimation of effect sizes in the original GWAS, which depends on the GWAS sample size, the ancestry composition, and the outcome definition used, and (ii) during the PRS construction step, where these GWAS estimates are subsequently selected, transformed, recalibrated, or combined using different statistical methods.

To distinguish between these competing explanations, we structured our analyses in a three-step framework. Step 1 evaluated whether the observed intra-individual variability is consistent with a shared underlying genetic signal subject to statistical noise, or with the capture of additional predictive information. Step 2 examined, under the statistical-uncertainty hypothesis, which factors drive the residual differences between PRSs, distinguishing the contribution of SNP inclusion (overlap and PRS size) from that of SNP weighting (GWAS source and PRS construction method). Step 3 then assessed whether grouping PRSs by shared construction features attenuated intra-individual variability, providing a practical framework to identify combinations of PRSs that yield more consistent individual risk estimates. Details on each step can be found in the Supplementary Methods.

### Clinical impact of PRS-related intra-individual variability after integration into SCORE2-OP

SCORE2-OP is the European Society of Cardiology-recommended tool for estimating cardiovascular risk in primary prevention.^17–19^ It classifies individuals into low, intermediate, and high risk using age-specific thresholds (Supplementary Methods). We constructed integrated scores by adding the PRS effect per 1-SD to the SCORE2-OP linear predictors, as previously done.^10,20,21^ We then recomputed population-level performance and intra-individual variability using guideline thresholds.

Finally, to provide a clinically accepted reference level of intra-individual variability against which the PRS-driven variability could be compared, we quantified how much SCORE2-OP estimates vary for the same individual when a clinical input is subject to its usual measurement error. We chose systolic blood pressure (SBP), which has large within-person variability (≈10 mmHg)^22,23^ For each participant, we drew 20 SBP values from a normal distribution (SD 10 mmHg) around the observed value and recomputed SCORE2/-OP.

## Results

### Sample characteristics

Of 4,137 participants (*Supplementary Figure 1*), 478 (12%) developed incident ASCVD over a median 14.4 years (IQR 10.7–14.6; incidence 9.3/1,000 person-years, 95% CI 8.5–10.2). Mean age was 53.5 ± 10.7 years, and 54% were female participants. Compared with individuals without ASCVD, those with ASCVD were older, more often male, and had a less favourable cardiometabolic and lipid profile and reduced kidney function (*Supplementary Table 3)*.

### Population-level performances of selected PRSs for ASCVD prediction

All 17 PRSs were associated with higher ASCVD risk, with a 1-SD increase corresponding to a 22–45% rise in risk. (*Supplementary Table 4*). PGS003725 served as reference (Brier 0.10, 95% CI 0.09–0.11; AUROC 0.59, 0.56–0.62) (*Supplementary Figure 2A and 2B*). All Brier differences fell within the ROPE, indicating equivalent calibration. For AUROC, all PRSs except PGS000012 had >95% of posterior samples within the ROPE (*Supplementary Table 5*). PGS000012 was therefore excluded, leaving 16 PRSs for the variability analysis (Table 1).

**Table 1.** Characteristics of PRSs assessed for individual-level variability in ASCVD risk estimates.

| PRS ID | Publication (DOI) | Source GWAS | Method for SNPs selection | Number of SNPs | GWAS ancestry |
| --- | --- | --- | --- | --- | --- |
| **External validation and added value compared to traditional scores** |  |  |  |  |  |
| PGS000013 | 10.1038/s41588-018-0183-z | CARDIoGRAMplusC4D 1000 Genomes-based GWAS | LDpred | 6,630,150 | Combined |
| PGS000018 | 10.1016/j.jacc.2018.07.079 | 1) new PRS from CARDIoGRAMplusC4D 1000 Genomes-based GWAS<br>2) PGS000012 score from CARDIoGRAMplusC4D Metabochip stage-2 meta-analysis<br>3) FDR202 score from CARDIoGRAMplusC4D 1000 Genomes-based GWAS | 1) C+T<br>2) C+T<br>3) GCTA (reference panel : reference panel of 1,000 genotyped sample, significant threshold : FDR P <0.05 ) | 1,745,179 | Combined |
| PGS000116 | 10.1001/jama.2019.22241 | CARDIoGRAMplusC4D 1000 Genomes-based GWAS | Lassosum | 40,079 | Combined |
| PGS000117 | 10.1001/jama.2019.22241 | CARDIoGRAMplusC4D 1000 Genomes-based GWAS | Lassosum | 297,862 | Combined |
| PGS001780 | 10.1038/s42003-021-02996-0 | CARDIoGRAMplusC4D 1000 Genomes-based GWAS | PRS-CS | 1,090,048 | Combined |
| PGS003438 | 10.1001/jamacardio.2022.4466 | Combined CARDIoGRAMplusC4D 1000 Genomes-based GWAS + CARDIoGRAM Exome + Howson's GWAS met-analysis + Webb's GWAS meta-analysis + 10 new studies (available in Table S3 in 10.1038/s41588-022-01233-6) | GCTA (reference panel : the UKBB sample using European ancestry participants only, significant threshold : Pjoint ≤ 5×10<sup>-8</sup> ) | 241 | >95% European |
| PGS003727 | 10.1038/s41591-023-02429-x | FinnGen\|CARDIoGRAMplusC4D without UKB\|MVP | LDpred2 | 1,125,113 | >95% European |
| PGS003726 | 10.1038/s41591-023-02429-x | European : FinnGen\|CARDIoGRAMplusC4D without UKB\|MVP : East Asian : BBJ; African : MVP; Hispanic : MVP; South asian : G&H | LDpred2 (**GPSMult** ; trait-specific combination) | 1,296,172 | Combined |
| PGS003725 | 10.1038/s41591-023-02429-x | European : FinnGen\|CARDIoGRAMplusC4D without UKB\|MVP : East Asian : BBJ; African : MVP; Hispanic : MVP; South asian : G&H | LDpred2 (**GPSMult+** ; cross-trait combination with risk factor)) | 1,296,172 | Combined |
| **New scores from 2024/2025 without external validation** |  |  |  |  |  |
| PGS004309 | 10.1038/s41467-024-48654-x | UK Biobank | GenoBoost | 3,000 | >95% European |
| PGS004696 | 10.1161/CIRCGEN.123.004272 | Combined BBJ\|CARDIoGRAMplusC4D\|MVP\|UKB | PRS-CSx | 1,289,980 | Combined |
| PGS004697 | 10.1161/CIRCGEN.123.004272 | Combined CARDIoGRAMplusC4D\|MVP\|UKB | PRS-CSx | 1,120,251 | Combined |
| PGS004698 | 10.1161/CIRCGEN.123.004272 | Combined BBJ\|CARDIoGRAMplusC4D\|MVP\|UKB | P+T | 542,218 | Combined |
| PGS004743 | 10.1016/j.xgen.2024.100523 | PGS Catalog | **PRSmix** (trait-specific combination) | 3,606,321 | Combined |
| PGS004745 | 10.1016/j.xgen.2024.100523 | PGS Catalog | **PRSmix+** (cross-trait combination with risk factors) | 4,769,577 | Combined |
| PGS004879 | 10.1038/s41467-024-48938-2 | CARDIoGRAMplusC4D 1000 Genomes-based GWAS | megaprs.auto | 610,677 | Combined |

### Individual-level variability in ASCVD risk prediction across the 16 practically equivalent PRSs

The median SD of risk percentiles across PRSs was 18.4 (95% CI 18.2–18.5). In practical terms, for a typical individual, estimates from different PRSs varied by roughly ±18 percentile points around their mean. The median CV was 0.42 (0.41–0.42), meaning the relative dispersion was about 40% of the individual’s mean percentile. (*Figure 1*). Agreement between PRSs was moderate (ICC 0.58, 0.56–0.59). Agreement in classifying individuals was also moderate: mean Cohen’s kappa was 0.41 (90th) and 0.37 (95th) for high-risk, and identical for low-risk (10th/5th).

**Figure 1.**
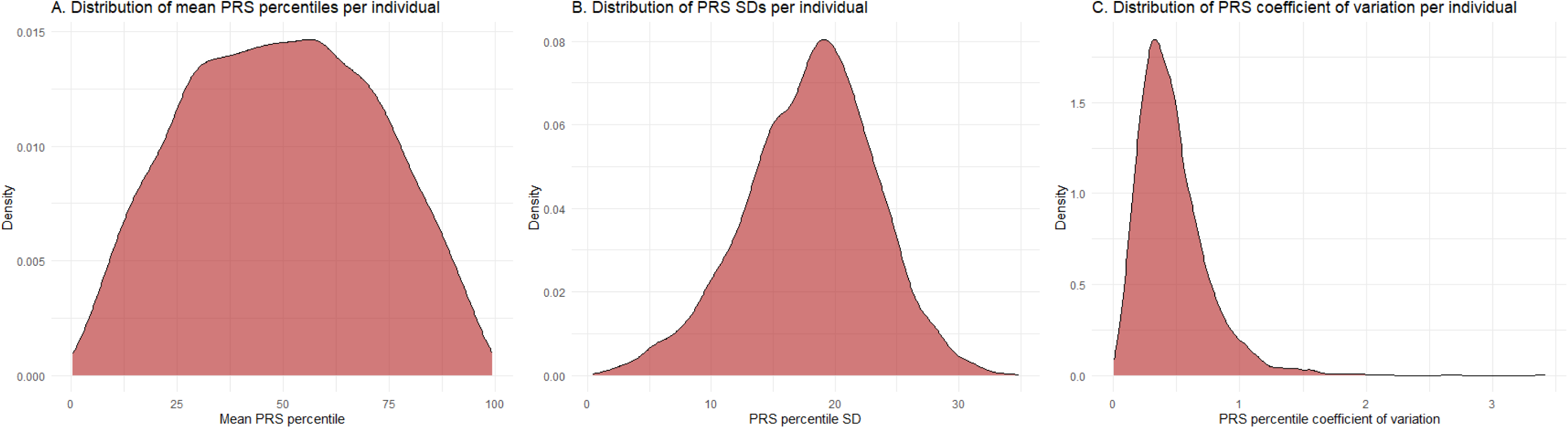
Distribution of individual-level mean ASCVD risk percentiles across polygenic risk scores. Panel A shows the distribution of mean ASCVD risk percentiles per individual, calculated by averaging the percentiles assigned by 16 distinct PRSs. This reflects the central tendency of polygenic risk-based classification for each participant. Panel B displays the distribution of within-individual standard deviations (SDs) of ASCVD percentiles, calculated as the square root of the average squared difference between each PRS-assigned percentile and the individual’s mean percentile and capturing the degree of disagreement among PRSs in estimating a participant’s risk. Panel C presents the distribution of individual-level coefficients of variation (CVs), defined as the ratio of the SD to the mean percentile, offering a scale-independent measure of variability. Together, these panels illustrate both the central tendency and variability in polygenic risk estimates at the individual level.

Overall, only a small proportion of individuals had PRS scores in both extremes simultaneously. 5.2% of participants had at least one score in both the top and bottom 10th percentiles. Among the 132 participants with more than two PRSs in the extreme percentiles, the PRSs most frequently observed in the opposite extreme relative to the majority were: PGS004309 (34 occurrences), PGS003438 (27), PGS000117 (16), PGS000116 (13), PGS004698 (13), PGS004697 (12), PGS004696 (7), PGS000018 (5), PGS004879 (5). Using the more stringent 5th and 95th percentile thresholds, falling to 0.87% at the more stringent 5th/95th percentiles (*Figure 2).* Among the 17 participants with more than two PRSs in the extreme percentiles, the PRSs most frequently observed in the opposite extreme relative to the majority were: PGS004309 (10), PGS003438 (3), PGS000117 (2). PGS003725 was the only PRS that showed no occurrences of discordance relative to the majority under either threshold.

**Figure 2.**
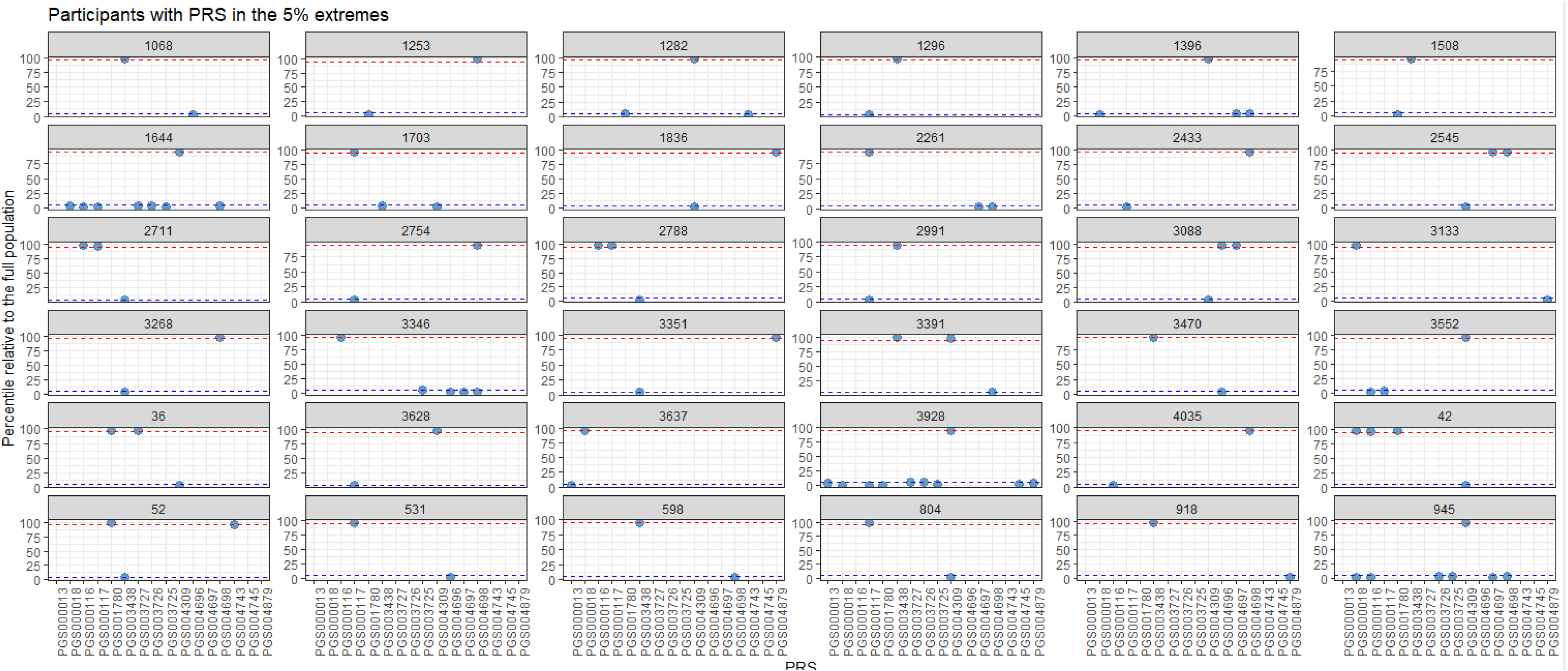
Participants with simultaneous high- and low-Risk classifications across multiple PRSs. The graph shows 30 participants with discordant extreme PRS values. Only extreme PRS estimates are shown. Specifically, for each individual, PRS values falling above the 95th percentile or below the 5th percentile of the distribution (x-axis) are displayed across the 16 PRSs (y-axis). PRS values between the 5th and 95th percentiles are not shown.

### Exploring sources of intra-individual variability across ASCVD-PRSs

#### Step 1 — Does variability reflect additional predictive information or statistical uncertainty?

To assess whether variability between PRSs reflects true additional predictive information or merely statistical uncertainty, we first compared the observed level of discordance with what would be expected by chance alone if all PRSs were measuring the same underlying genetic risk, estimated here using Monte Carlo simulations that preserved the correlation structure between PRSs. The observed proportion of individuals classified in opposite extremes (top and bottom percentiles) closely matched the proportions expected under this model. At the 10th/90th percentiles, 5.2% were discordant versus 5.5% expected (95% simulation interval 4.8–6.2%). At the 5th/95th percentiles, 0.87% versus 0.84% expected (0.56–1.11%). The close match suggests the disagreement reflects statistical variability, not systematic additional information. We further examined whether individuals with discordant PRS classifications displayed specific phenotypic characteristics that might suggest distinct biological mechanisms captured. No consistent differences were observed in cardiovascular risk factors, lifestyle, mental health, or ASCVD outcomes between individuals with and without discordant PRSs (*Supplementary Table 6 and Table 7*). The only exception was elevated total cholesterol (>7 mmol/L). Yet if this reflected biology captured by specific scores, the PRSs incorporating risk-factor GWAS (PGS003725, PGS004745) should classify these individuals as high-risk more often, and they did not. This absence of a phenotypic signature argues against complementary information.

Across full variant sets, overlap, measured by the Jaccard index, the fraction of SNPs shared between two scores, was low (median Jaccard 0.04, IQR 0.006–0.20; Supplementary Table 8). This low global overlap, however, masked a different pattern. The median proportion of shared SNPs relative to each individual score was higher (34.5%, IQR 12.5–52.3%), meaning smaller PRSs often share much of their content with larger ones. Consistent with this, ≥70% of the smaller PRS’s SNPs were nested within the larger PRS in 35% of comparisons (Supplementary Figure 3). These nested relationships formed a hierarchy, with intermediate PRSs (PGS003726, PGS003725, PGS003727) connecting smaller scores to the largest (PGS000013). Low Jaccard values and strong nesting are therefore not contradictory: they reflect differences in PRS size around a common genetic core. This is captured by the SNP count ratio, the ratio of the number of SNPs in the two scores, which reflects how comparable they are in size, which was low (median 0.23, IQR 0.002–0.49), confirming that pairs often combined a much smaller score with a much larger one. Despite this limited overlap, PRSs remained moderately correlated at the individual level (mean r ≈ 0.54). Had PRS-specific SNPs captured distinct information, low-overlap pairs would have ranked individuals very differently; instead, the persistence of moderate correlation points to a largely shared underlying signal, making it unlikely that the variability is driven primarily by additional risk information.

Finally, we tested directly whether PRS-specific variants add predictive value. We decomposed each pair into shared and unique components by linkage disequilibrium (LD) structure, then modelled their contributions (*Supplementary Table 9)*. The shared component consistently predicted ASCVD, whereas unique components contributed less often and less strongly (*Supplementary Figure 4*). Only 28 of 120 comparisons showed a significant unique contribution after FDR correction and accounting for multicollinearity (Figure 3; *Supplementary Table 10*; *Supplementary Figure 5)*. Some PRSs (PGS004697, PGS004696, PGS003727, PGS003726, PGS003725) showed unique components more frequently (*Supplementary Figure 6*). Moreover, shared components were moderately-to-highly correlated across individuals (median 0.67, IQR 0.59–0.75), meaning PRSs rank individuals similarly once restricted to shared information (Figure 3). Together, these findings suggest that most PRSs capture a largely common genetic signal, with limited additional predictive value from PRS-specific variants.

**Figure 3.**
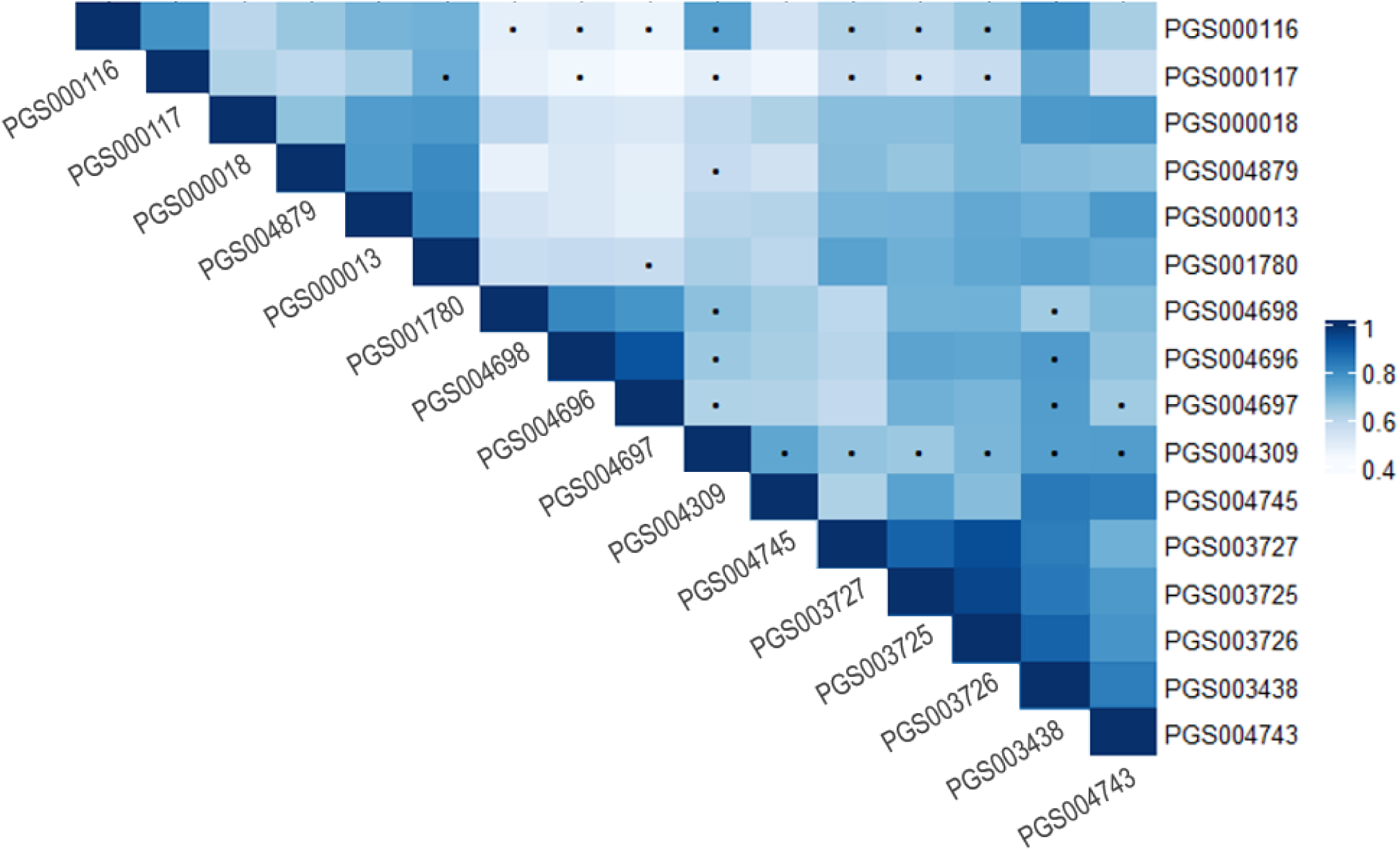
Overlap-score concordance and independent predictive contribution across ASCVD polygenic risk scores. Heatmap showing Pearson correlation coefficients (r) between the *overlap components* of each pair of ASCVD-PRSs, reflecting the degree of shared genetic signal after LD-based pruning. Darker colors indicate stronger concordance and greater redundancy in captured genetic risk, whereas lighter colors indicate weaker concordance and more distinct genetic architecture. Cells marked with a dot (•) denote PRS pairs for which at least one *unique component* remained significantly associated with CVD status in the joint logistic regression model after false discovery rate (FDR < 0.05) correction, indicating the presence of PRS-specific predictive information beyond the shared genetic signal.

#### Step 2 — What drives intra-individual instability in PRS construction?

Having found the variability consistent with statistical uncertainty, we sought its drivers. Greater SNP overlap was associated with more similar estimates (p < 0.001) but explained only a modest share of the variability (R² ≈ 0.29), suggesting that low SNP overlap alone is not the primary driver. We therefore examined whether the SNP count ratio, the similarity in size between two scores, regardless of how many SNPs they share, could better explain this variability. PRSs of more comparable size produced more similar rankings (p < 0.001). When overlap and size were modelled together, only size remained independently associated with ranking differences (p = 0.003), indicating that differences in the number of variants included play a more important role than exact SNP overlap.

SNP weighting, and therefore model construction, also appeared to play a role, as PRSs with identical or highly overlapping SNP sets could still produce different individual risk estimates. Two PRSs with identical SNP sets (PGS003725/PGS003726, Jaccard 1) still differed by ∼7 percentile points despite a high correlation (r = 0.95) and no individuals were classified at opposite extremes. Across six pairs with Jaccard > 0.9, differences of ∼5–10 points persisted despite r > 0.9, again with extreme discordance rare or absent. Together, these findings indicate that intra-individual variability between PRSs is primarily driven by differences in scale (i.e., the number of SNPs included) and by how SNP effects are weighted, rather than by the inclusion of entirely distinct sets of genetic variants. Additionally, we added differences in how SNP effects are estimated and combined during PRS construction to the model, on top of SNP overlap and SNP count ratio. Several construction-related features were associated with reduced ranking variability and fewer extreme discordances. PRSs with similar ancestry composition (European-only or multi-ancestry) and the same GWAS composition category (CARDIoGRAM-only, CARDIoGRAM-based combinations, or PRS-specific GWAS sources) showed both reduced ranking differences (FDR-adjusted p = 0.02 and p < 0.0001, respectively) and fewer extreme discordances (FDR-adjusted p = 0.0005 and p = 0.04, respectively). Ensemble-based pairs, that is, PRSs that combine several existing PRSs, whether from different GWAS for the same trait, from different ancestries, or from genetically correlated traits, into a single integrated score, also showed reduced ranking differences (p = 0.004). In contrast, sharing the same baseline construction method was not associated with variability. Together, these results indicate that while SNP overlap and SNP count ratio contribute to inter-PRS differences in individual rankings, disagreement between PRSs, particularly at the extremes, is also shaped by how genetic effects are estimated in the source GWAS and how they are subsequently combined during PRS construction.

#### Step 3 — Does grouping PRSs by construction features reduce individual-level variability?

We grouped PRSs by shared features (Table 1) and reassessed variability (*Supplementary Table 11*). Grouping by GWAS source or ancestry resulted in only modest reductions in variability. Grouping by construction characteristics gave larger ones, particularly Bayesian methods, ensemble approaches, and large SNP counts. When we restricted the 9 PRSs with similar construction features (Bayesian-based methods with or without ensemble-based approaches and ≥1 million SNPs), both variability and extreme discordance decreased. Together, these findings suggest that selecting PRSs with similar construction characteristics can meaningfully improve the consistency of individual risk estimates and further support the conclusion that intra-individual variability is driven primarily by methodological differences in PRS construction rather than by complementary risk information.

### Comparison of individual-level variability in ASCVD-PRSs integrated into SCORE2

We then integrated the 16 PRSs into SCORE2-OP. The median SD of percentiles was 16.5 (95% CI 16.2–16.9) and the median CV 0.37 (0.37–0.38), with moderate agreement (ICC 0.53, 0.52–0.55). The continuous estimates were thus slightly more stable than the PRSs alone. The categorical picture, however, was very different. Mean weighted Cohen’s kappa across categorical models was 0.29, only fair agreement, indicating substantial inconsistency, including occasional large category (i.e., low to high) shifts for the same individual (*Supplementary Figure 7 for pairwise details)*.

Across models, 75.6% of participants were placed in different categories by at least one model. In practical terms, for approximately three out of four participants, at least one model placed them in a higher or lower risk group compared with another model. Moreover, 54.6% were classified as both low and high risk across models, showing that extreme reclassification was not uncommon. Categorical instability was concentrated among participants sitting close to a guideline threshold: those whose classification changed across models had a significantly smaller minimum distance to the nearest cut-point than those who remained stable (*Supplementary Figure 8*; p < 0.001).

Using SCORE2-OP + PGS004745 as reference, given its highest discrimination (AUROC 0.76, 0.74–0.79), agreement varied sharply by category (Figure 4). Among participants classified as low risk by the reference model, most participant–model comparisons remained low risk across the other models (83.1%), whereas 10.1% were reclassified as intermediate risk and 6.8% as high risk. In contrast, classification was less robust among participants classified as intermediate or high risk by the reference model. Among those classified as intermediate risk, only 37.5% of comparisons remained intermediate, while 38.9% were reclassified as low risk and 23.6% as high risk by the other models. Similarly, among those classified as high risk by the reference model, 53.4% of comparisons remained high risk, whereas 23.2% shifted to intermediate risk and 23.4% to low risk. These findings indicate that PRS-integrated risk classification is relatively stable, in terms of intra-individual variability, for individuals classified as low risk, but substantially more sensitive to the choice of PRS among those classified as intermediate or high risk by the reference model.

**Figure 4.**
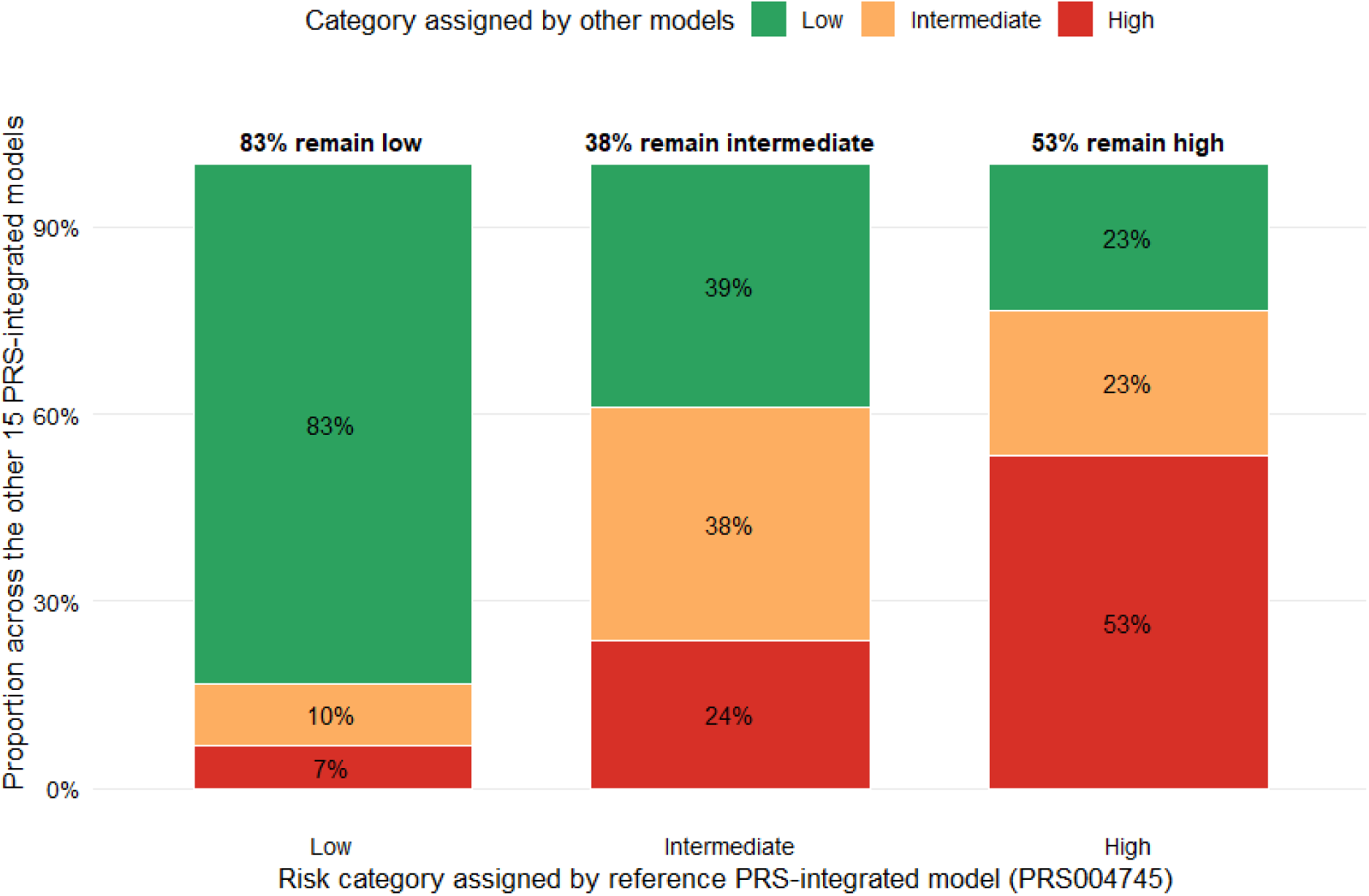
Intra-individual stability of PRS-integrated risk classification across models. Each bar represents individuals classified into a given risk category by the reference model (SCORE2-OP + PRS004745). Colors indicate how these same individuals are classified by the other 15 PRS-integrated models. A dominant single color reflects consistent classification across models, whereas a heterogeneous distribution indicates greater intra-individual variability.

Finally, we benchmarked this variability against a common real-world source of uncertainty: variation in SBP measurements. Using 21 SBP values per individual (20 simulated with measurement error, plus the observed value), variability was much lower than across the PRS-integrated scores: median SD 3.5 (95% CI 3.4–3.5, i.e. ≈ ±3 percentile points), median CV 0.09, and ICC 0.98. Categorical agreement also remained high (mean weighted Cohen’s kappa 0.82): 23.4% of individuals were reclassified by at least one model, but none shifted between low and high risk. The variability introduced by the choice of PRS therefore exceeds that of realistic measurement error in an established clinical input.

## Discussion

We compared a large set of ASCVD-PRSs with practically equivalent population-level performance and asked a simple question: do scores that look interchangeable at the population level agree when applied to the same person? They frequently did not. For a typical participant, genetic risk estimates differed by roughly ±18 percentile points across scores, and this disagreement persisted, and translated into unstable clinical categories. once the scores were embedded in the SCORE2-OP model. Crucially, this variability arose largely from statistical and methodological uncertainty rather than from the capture of additional or complementary risk information.

### Population-level equivalence does not guarantee individual-level agreement

Among PRSs that were statistically equivalent in overall discrimination and calibration, individual rankings still diverged substantially (ICC 0.58; moderate Cohen’s κ). This mirrors prior methodological work: Chen et al. showed that CHD-PRSs derived from different imputation pipelines are highly correlated overall yet can differ by more than 20 percentile points for individual patients,^24^ and Nguyen et al. reported correlations above 0.97 between array- and sequencing-based scores alongside non-negligible individual discrepancies.^25^ Our results also closely align with those of Abramowitz et al.^13^, who evaluated 46 CHD-PRSs with practically equivalent population-level performance and reported a similar magnitude of intra-individual variability. Notably, only five PRSs overlapped with our study, suggesting that this variability reflects a general property of PRS-based risk estimation rather than specific score selection. Despite these similarities, we observed substantially less discordance at the extremes of the risk distribution. Using the 5th and 95th percentile thresholds, fewer than 1% of participants in our cohort showed opposing classifications across PRSs, compared with approximately 20% in the Abramowitz study. Discordance remained lower in our data even at the 10th and 90th percentile thresholds (5.2%).

### Variability reflects estimator uncertainty rather than capturing genetic signal

Our second aim was to weigh two possible interpretations of the disagreement between scores. Under the first, the distinct SNPs in each PRS would capture complementary risk information, so the variability itself is informative; under the second, these SNPs would simply represent different statistical attempts to capture the same underlying signal, so the variability reflects noise rather than information. Three converging findings favour the second. First, the frequency of individuals at opposite risk extremes matched what chance alone would produce if all PRSs measured a single shared signal with random noise, with no enrichment of discordant individuals and no distinct clinical or outcome profile among them. Second, despite sharing only a small fraction of their SNPs, the scores remained moderately correlated at the individual level, a pattern that would be difficult to reconcile with PRSs capturing genuinely different information. Third, when each pair of scores was decomposed into shared and score-specific components, most of the predictive power appeared to reside in the shared component, with score-specific effects reaching significance in only a minority of comparisons.

This statistical noise can arise at the two levels inherent to any PRS. The first is the limited overlap in SNP composition: scores tag the same signal through different, imperfectly interchangeable variants. This, however, did not appear to be the principal driver, once score size was accounted for, exact overlap was no longer independently associated with disagreement. The second level, which seemed to contribute more, concerns the SNP weights: scores with near-identical SNP sets could still differ by several percentile points per individual, and this weighting-related variability tended to track both the GWAS source (ancestry, provenance) and the construction step (score size, Bayesian shrinkage, ensemble methods). A comparable mechanism has been described at the level of a single score by Ding et al., who showed that finite GWAS sample size and LD make the assignment of effects among correlated variants inherently uncertain, so that an individual’s estimated genetic risk carries a large statistical variance, an individual at the 90th percentile could plausibly lie anywhere from the 34th to the 99th once this uncertainty is accounted for.^11^ The disagreement we observe between equivalent scores may therefore largely represent the between-PRS expression of the same estimation uncertainty they describe within a single score. Taken together, these observations suggest that most PRSs may be capturing largely the same genetic risk through imperfect proxies, differing mainly in how that shared signal is estimated and weighted rather than in the information they capture.

### Construction choices modulate, but do not eliminate, discordance

PRS construction also influenced this variability. Scores that included more than one million variants and those built with Bayesian or ensemble-based methods showed lower intra-individual variability. This fits the principle illustrated by Monti et al., who benchmarked seven construction methods across five biobanks: no single method was consistently best, but combining several methods into an ensemble gave the most consistent and transferable results.^26^ In both cases, combining information, across many variants or across several methods, reduces the influence of any single construction choice and produces more stable estimates. Our findings therefore point to two complementary levers for improving intra-individual consistency: harmonising construction methods and favouring PRSs that retain a large number of variants rather than restricted variant sets. These measures may reduce, but not eliminate, the variability between scores, an important consideration for implementation, since this uncertainty becomes clinically meaningful once continuous estimates are turned into treatment-guiding categories.

### Integration into clinical risk model amplifies categorical instability

Integrating PRSs into SCORE2-OP slightly improved the stability of continuous estimates but markedly destabilised categorical classification: about three in four participants were placed in different risk categories depending on which equivalent PRS was used, and more than half were classified as both low and high risk across models. This apparent paradox, more agreement on a continuous scale, yet frequent category switches, is explained by how close much of the cohort sits to a guideline threshold. Measuring each participant’s absolute SCORE2-OP risk against the age-specific cut-points that define the low, intermediate, and high categories, we found the median distance to the nearest threshold was only 1.9 percentage points; 26% of participant–model estimates fell within 1 percentage point of a cut-point, and 57% within 2 percentage points. Consistent with this, participants who changed category across models lay closer to a threshold than those whose classification remained stable. This instability was concentrated among individuals labelled intermediate or high risk by the reference model, whereas low-risk classifications were comparatively robust. To benchmark this against uncertainty already accepted in practice, we let SCORE2-OP vary with realistic measurement error in systolic blood pressure: agreement remained very good overall, although 23.4% participants changed risk category versus 75.6% across PRS models. Importantly, no patient shifted between low and high risk. PRS-related uncertainty thus substantially exceeds that of a routinely tolerated clinical input, an important caveat to studies reporting average reclassification gains^1^, which do not capture sensitivity to PRS choice at the individual level. In practice, this means that treatment eligibility, intensity of preventive interventions, and patient counseling may differ substantially depending on which PRS is integrated, raising concerns about clinical standardization.

### Implications for clinical deployment

These findings suggest that PRSs remain non-interchangeable clinical tools: “adding a PRS” to a risk model is not a well-defined intervention without prespecified scores, calibration procedures, and disclosure frameworks. Although such variability may be acceptable for population-level screening, divergent individual-level reliability complicates personalised treatment decisions, because the choice of PRS, rather than the patient’s underlying risk profile, can influence the recommendation. Although large, Bayesian, or ensemble-based methods improve stability, methodological standardisation remains necessary.

A conservative implementation strategy would position PRSs as secondary risk enhancers rather than as interchangeable primary terms in clinical risk equations. In this role, PRSs could act as tie-breakers for borderline or intermediate-risk individuals, where modest shifts in estimated risk may plausibly alter management while limiting the clinical consequences of estimator uncertainty. This targeted approach is consistent with modelling evidence suggesting that PRS-informed strategies may be most efficient in intermediate-risk strata and that targeted re-screening of individuals at intermediate SCORE2 risk may prevent more CVD events than broader population-wide implementation.^27,28^ For individuals already at high clinical risk, PRSs should not be used to recalculate risk in a way that overrides established indications. However, standardized disclosure may still have a complementary role in risk communication and shared decision-making, as observational and implementation studies suggest that genetic risk disclosure can improve understanding and, in some settings, support preventive behaviour or management changes, although a recent systematic review indicate that these effects are modest and heterogeneous overall.^29–31^

Ultimately, residual individual-level uncertainty reinforces the need to evaluate PRS disclosure prospectively rather than infer clinical utility from retrospective discrimination or reclassification metrics alone. Pragmatic trials embedded within existing clinical infrastructures are required to determine whether standardized PRS selection and disclosure improve preventive treatment uptake, patient understanding, risk-factor control, and long-term cardiovascular outcomes.

## Strengths and limitations

Strengths include a head-to-head comparison of a large PRS panel in a well-characterised cohort. Importantly, this comparison was performed in a cohort that was used neither to derive nor to validate any of the scores. This is a meaningful advantage: most of the PRSs we evaluated were developed and tested in large biobanks, particularly the UK Biobank, and a score generally performs better, and more stably, in the very sample used to build or tune it. Evaluating the scores in an external, independent population therefore provides a more realistic picture of how they would behave in routine clinical use, and makes it unlikely that the intra-individual variability we observed is an artefact of overlap between our cohort and the scores’ training or validation data.

Several limitations warrant consideration. First, our sample was of predominantly European ancestry, in which discordance between scores are likely to be smaller than in more diverse populations, where PRS transferability is lower.^32^ Second, our central question, whether the disagreement between scores reflects genuinely additional genetic information or merely statistical noise, could only be addressed indirectly. Distinguishing these possibilities directly would require quantifying the statistical uncertainty attached to each SNP’s effect, for example through the individual-level credible intervals proposed by Ding et al.^11^, which in turn requires multivariable SNP standard errors that are not currently available in the PGS Catalog. Finally, our analysis examined a single outcome (ASCVD) within a single clinical framework (SCORE2-OP); whether comparable intra-individual variability and categorical instability arise for other diseases or other risk models remains to be established.

## Conclusions

ASCVD-PRSs that are practically equivalent at the population level can still yield substantially different risk estimates, and different clinical classifications, for the same individual. This disagreement appears to arise mainly from how a shared genetic signal is estimated and weighted across scores, rather than from genuinely complementary risk information. Construction choices, notably the number of variants and the use of Bayesian or ensemble-based methods, can attenuate but not remove it. After integration into SCORE2-OP, classification stayed stable for low-risk individuals but was sensitive to the choice of score at intermediate or high risk, where it can change management. Overall, equivalent population-level performance is not sufficient to treat PRSs as interchangeable at the individual level, and methodological standardisation and pragmatic clinical trials remain necessary to determine whether PRS integration improves long-term cardiovascular outcomes.

## Acknowledgements

The authors would like to thank all the people who participated in the recruitment of the participants, data collection and validation, particularly Nicole Bonvin, Yolande Barreau, Mathieu Firmann, François Bastardot, Julien Vaucher, Panagiotis Antiochos, Cédric Gubelmann, Marylène Bay, Benoît Delabays and Adelin Barrier.

## Data availability statement

The data of CoLaus|PsyCoLaus study used in this article cannot be fully shared as they contain potentially sensitive personal information on participants. According to the Ethics Committee for Research of the Canton of Vaud, sharing these data would be a violation of the Swiss legislation with respect to privacy protection. However, coded individual-level data that do not allow researchers to identify participants are available upon request to researchers who meet the criteria for data sharing of the CoLaus|PsyCoLaus Datacenter (CHUV, Lausanne, Switzerland). Any researcher affiliated to a public or private research institution who complies with the CoLaus|PsyCoLaus standards can submit a research application to. Proposals will be evaluated by the Scientific Committee (SC) of the CoLaus|PsyCoLaus study. Detailed instructions for gaining access to the CoLaus|PsyCoLaus data used in this study are available at www.colaus-psycolaus.ch/professionals/how-to-collaborate/.

