## Supplementary Methods and Figures for "Advancing Clinical Implementation of Cardiovascular Polygenic Risk Scores Through Patient-Level Robustness Assessment"

### Supplementary Material

### Supplementary Methods

#### *Single-Nucleotide-Polymorphisms (SNPs) genotyping in CoLaus|PsyCoLaus*

DNA samples from 5,399 participants were genotyped for 799,653 SNPs using the BB2 GSK-customized Affymetrix Axiom Biobank array. Quality control procedures, as described by Anderson et al., were implemented.[1]

There were no exclusions due to missing SNP rates greater than 5%. Participants whose genetic sex did not match their self-reported gender were removed from the study.

After screening with Kinship-based Inference for Genome-wide association studies (KING v2.2.6), no participants were found to be related.

Additionally, Principal Component Analysis (PCA) was conducted using PLINK (v2.00a3LM) in conjunction with the 1000 Genomes Project reference panel, and no participants of non-European ancestry were identified for exclusion.

Subsequently, SNPs were excluded if they were missing in more than 5% of the samples or if they deviated significantly from Hardy-Weinberg equilibrium (HWE, P<10e-7). This process resulted in the retention of 697,923 SNPs among 4'781 participants for haplotype determination and imputation. To statistically estimate the haplotypes, the genotypes were phased using EAGLE2 v2.0.5 software and the Haplotype Reference Consortium (HRC) r1.1 reference panel.[2–4]

Genotype imputation was performed using two independent reference panels: the HRC reference panel and the merged 1000 Genomes Phase 3 and UK10K reference panel.[3,5,6] Both phasing and imputation were performed on Sanger Imputation Service. The imputed data set (N=89 million SNPs) consisted of a) SNPs imputed with HRC reference panel only, b) SNPs imputed with the merged reference panel only, and c) SNPs with the highest imputation information (INFO) score if imputed with both reference panels. To filter out poorly imputed or rare genotypes, SNPs with INFO score < 0.8, minor allele frequency <1% or significant deviation from HWE were excluded, resulting in a final number of 9'031'264 SNPs available for PRS calculation.

#### *PRS calculations*

To compute the PRSs, we used the scoring files available in the PGS Catalog, which include the relevant SNPs, their effect alleles, and corresponding effect sizes derived from relevant GWAS. Each SNP is assigned a weight based on its effect size from the GWAS. The weight reflects the strength of the association between the SNP and the disease. he PRS is calculated by summing the weighted effects of the selected SNPs. This can be done using the formula:

$PRS=\sum( w_i \times g_i )$, where $w_i$ is the weight (effect size) of SNP i and $g_i$ is the genotype of SNP i (usually coded as 0, 1, or 2, depending on the number of risk alleles among the two haplotypes). This approach allows us to capture the cumulative genetic risk associated with multiple variants.

To ensure consistency and comparability across the PRSs and within the clinical risk model, the PRS values were normalized by converting them into z-scores. This process involves subtracting the mean of the PRS values from each individual PRS value and then dividing by the standard deviation. As a result, the z-scores represent how many standard deviations each PRS value is from the mean, allowing for direct comparison across different PRSs on a standardized scale.

Following normalization, participants were categorized into two risk groups based on the distribution of PRS values. The high-PRS risk category included participants in the top ≥90th percentile or the top ≥95th percentile of the PRS distribution for the different analysis.

***Bayesian assessment of population-level equivalence***

PRSs were first filtered to retain only those with equivalent population-level performance. This step was essential to ensure that any subsequent differences observed at the individual level would reflect differences in risk distribution rather than disparities in overall predictive accuracy. As an initial quality control step, the 17 selected and normalized PRSs were evaluated using Cox proportional hazards models to assess their association with incident ASCVD in the CoLaus|PsyCoLaus longitudinal cohort. The follow-up period for each participant was calculated from the baseline assessment to the earliest of the following censoring events: the development of the outcome, the last follow-up contacts if there was no non-cardiovascular death thereafter, or non-cardiovascular death. The models were adjusted for age, sex, and the first 10 principal components to account for population stratification. PRSs that did not show a nominally significant association with ASCVD (p-value (*p*) > 0.05) were excluded from further analyses. We then compared their predictive performance using a Bayesian analysis of variance approach in R, as described by Abramowitz and colleagues[7]: for each PRS, predictive performance was estimated using the Brier score and the area under the receiver operating characteristic curve (AUROC) from logistic regression models. Uncertainty was quantified via bootstrapped 95% confidence intervals. Pairwise differences in Brier scores and AUROCs relative to a reference PRS (lowest Brier score and/or highest AUROC) were modeled using Bayesian posterior distributions. A region of practical equivalence (ROPE) of ±0.02 was applied, and PRSs with ≥95% of posterior samples within the ROPE were considered statistically equivalent to the reference.

***Detailed metrics of intra-individual variability***

For each participant, we first computed a mean risk percentile by averaging the percentiles assigned by all retained PRSs. Within-person variability was then summarised with two complementary measures.

The standard deviation (SD) was calculated as the square root of the average squared difference between each PRS-assigned percentile and the individual's mean percentile. The coefficient of variation (CV) was defined as the SD divided by the mean percentile. When all PRSs assigned the same percentile to an individual, both SD and CV equalled zero, indicating no variability; higher values indicated greater disagreement among PRSs. Bootstrapped 95% confidence intervals for the median SD and CV were obtained using a nonparametric bootstrap with 1,000 resamples in base R.

We computed the intraclass correlation coefficient (ICC) to assess the consistency and precision of risk estimates across the different PRSs, treated as raters. ICC values were interpreted as poor (<0.5), moderate (0.5–0.75), good (0.75–0.9), or excellent (>0.9) reliability.[8]

Agreement in categorical classification was assessed with Light's kappa, applied to high-risk classification (above the 90th or 95th percentile) and low-risk classification (below the 10th or 5th percentile).[9,10] We additionally computed the proportion of individuals whose PRSs fell simultaneously into a high and a low percentile category. Finally, among individuals with at least three PRSs in an extreme decile (top or bottom), we identified PRSs falling in the opposite extreme relative to the majority of that individual's extreme scores, and quantified how often each PRS appeared as an "opposed" score across participants, to flag PRSs contributing more strongly to intra-individual variability.

#### *SCORE2-OP risk categories and thresholds*

ESC guidelines recommend classifying individuals into three age-specific cardiovascular risk categories using SCORE2-OP. [11]

| **Risk category** | **<50 years** | **50–69 years** | **≥70 years** | **Recommended action** |
|---|---|---|---|---|
| Low | <2.5% | <5% | <7.5% | Lifestyle modification; no pharmacological treatment |
| Intermediate | 2.5–7.5% | 5–10% | 7.5–15% | Preventive treatment to be considered |
| High | ≥7.5% | ≥10% | ≥15% | Preventive treatment recommended |

Additional details on the implementation of SCORE2-OP in the CoLaus|PsyCoLaus cohort are provided here.[12]

#### *Different methodological approaches to polygenic risk score construction*[13–15]

The construction of a polygenic risk score (PRS) typically relies on two distinct datasets and proceeds in several stages. The first input is a set of GWAS summary statistics — most importantly, the estimated effect size and p-value associated with each single nucleotide polymorphism (SNP) — which provides the raw material for identifying variants potentially linked to the trait of interest. However, these summary statistics alone do not indicate how well a given combination of variants will actually predict the trait in a new individual. To address this, a second, independent dataset is required: a development (or training) dataset containing individual-level genotype and phenotype data. This dataset is used to tune the parameters of the PRS — such as the p-value threshold for including SNPs, or the way correlations between nearby variants (linkage disequilibrium, or LD) are handled — in order to identify the configuration of SNPs and effect-size weights that yields the best predictive performance. Once this optimal configuration has been determined, the resulting score can then be evaluated in a third, fully independent testing dataset.

Many statistical methods have been developed to build a PRS. All of them produce, in the end, the same kind of output: a list of selected genetic variants (SNPs), each associated with a numerical weight. An individual's PRS is then simply the sum of the risk alleles they carry, each multiplied by its weight.

Where these methods differ is in how they decide which variants to include and how they compute each variant's weight. Two main challenges shape these choices:

- Nearby variants tend to be inherited together — a phenomenon called linkage disequilibrium (LD) — which means their GWAS effect sizes are not independent and tend to be inflated for variants that "tag" a true causal signal.
- The raw effect sizes reported by a GWAS are noisy estimates, especially for variants with weak signals, and using them directly tends to produce inaccurate predictions.

The methods below address these challenges in different ways. They fall into four broad families: (i) LD-based filtering and thresholding methods, (ii) Bayesian methods, (iii) ensemble-based approaches, and (iv) other approaches.

#### *(i) LD-Based Filtering & Thresholding Methods*

These methods reduce the noise in two steps: they first remove redundant variants (those carrying overlapping information due to LD), then keep only those most strongly associated with the trait. The weights themselves are not recomputed: they are taken directly from the GWAS summary statistics (the per-allele effect size β, or the log odds-ratio for binary traits).

##### Genome-wide Complex Trait Analysis (GCTA)

This method identifies variants that each contribute *independently* to the trait. In each genomic region, it starts from the variant with the strongest GWAS signal, then asks: once this variant is statistically accounted for, do any nearby variants still show a significant association? If so, they are also retained. This is done through a *conditional and joint regression* using GWAS summary statistics and an LD reference panel. Final weights are the joint regression coefficients — that is, the GWAS effect sizes re-estimated *jointly* for the selected independent variants, so that each weight reflects that variant's own contribution rather than a signal partly borrowed from a neighbour.

##### Pruning and Threshold (P +T)

The *pruning* step removes variants that are too correlated with one another (typically $r^2 \geq 0.1$ within a 250 kb window), keeping only one representative per correlated group — chosen without regard to its p-value. The *thresholding* step then keeps only variants with a GWAS p-value below a chosen cut-off. Both thresholds are tuned on the development dataset. Final weights are the original GWAS effect sizes of the selected variants, used as-is.

Clumping and Threshold (C+T)

C+T pursues the same goal as P+T but uses a smarter procedure. Instead of pruning variants without considering their significance, it starts from the *most significant* variant in each region and groups ("clumps") together all nearby variants in high LD with it (e.g. $r^2 \geq 0.1$ within 250 kb), keeping only the lead one. The procedure is then repeated with the next most significant remaining variant. A p-value threshold is then applied, and both the LD and p-value thresholds are tuned on the development dataset. As with P+T, final weights are the GWAS effect sizes of the retained lead variants, used unchanged

#### *(ii) Bayesian LD-Adjusted Effect Size Estimation*

Filtering methods keep only a small subset of variants and use their GWAS effect sizes directly. Bayesian methods take a fundamentally different approach: they keep (almost) all variants and recompute their weights.

The idea is that the raw GWAS effect sizes are noisy and inflated by LD, so they should not be used as-is. Instead, Bayesian methods combine two sources of information: (i) the GWAS summary statistics, and (ii) a *prior* assumption about how SNP effect sizes are distributed across the genome (for example, "most variants have no effect, a few have moderate effects"). The result, for each variant, is a posterior mean effect size — essentially, a refined estimate that is "pulled" toward zero (shrunk) when the GWAS signal is weak, and barely modified when the signal is strong. LD information from a reference panel is used so that this shrinkage properly accounts for the fact that correlated variants share part of their signal.

An important point about SNP selection. Bayesian methods do not typically operate on the entire set of variants reported by the GWAS. In practice, they are applied to a pre-defined panel of variants selected for their imputation quality and allele frequency, intersected with the variants available in the GWAS summary statistics. The size of this panel varies considerably across studies — from around 1 million variants (e.g. the HapMap3 panel, historically the most common choice) to several million when using denser panels such as 1000 Genomes, TOPMed, or UK Biobank imputed data. Within this input panel, *all* variants receive a non-zero weight: the model does not exclude any of them, but assigns very small weights to those whose signal is weak.

Linkage Disequilibrium-adjusted Polygenic Risk Score (LDpred)

LDpred assumes that only a fraction $p$ of variants are truly causal (e.g. $p = 0.001$, meaning 0.1% of variants), and that the rest have no effect. Using this assumption together with GWAS summary statistics and an LD reference panel, the method computes, for each variant, a posterior mean effect size that simultaneously (i) shrinks weak signals toward zero and (ii) deflates the effect sizes of variants that owe part of their GWAS signal to LD with a true causal variant. The value of *p* is tuned on the development dataset, and the posterior effect sizes obtained under the best-performing *p* are the final weights.

LDpred2

LDpred2 uses the same Bayesian principle as LDpred but introduces several improvements. It is faster, handles LD more accurately (using sparse LD matrices), and does not require the user to guess in advance the fraction *p* of causal variants. Instead, it tests a *grid* of plausible values for *p* (from 0.0001 to 1) and for the heritability scale, computes posterior mean effect sizes under each setting, and selects the combination giving the best predictive accuracy on the development dataset. The weights used in the final score are the posterior mean effect sizes obtained under this best-performing configuration.

PRS-Continuous Shrinkage (PRS-CS)

PRS-CS uses a different type of prior, called a *continuous shrinkage* prior. Rather than assuming that variants either have an effect or none at all (as LDpred does), it assumes a smooth distribution in which most variants have very small effects and a few have larger ones. The method then computes, for each variant, a posterior mean effect size that is gradually shrunk toward zero — weak signals are heavily shrunk (almost to zero), while strong signals are barely affected. A single parameter $\phi$ controls the overall amount of shrinkage; it can either be tuned on the development dataset or estimated automatically from the GWAS summary statistics. These posterior effect sizes serve as the final weights.

PRS-CSx

PRS-CSx extends PRS-CS to handle GWAS data from several ancestries simultaneously. It computes posterior mean effect sizes *jointly* across populations, using a shared continuous shrinkage prior that links effect sizes across ancestries while allowing them to differ in magnitude. This produces one set of weights per population; these per-population weights are then combined linearly into a single final score, with combination coefficients optimized on the development dataset. This approach improves prediction in non-European and admixed individuals, who are typically underserved by PRSs derived from European-only GWAS.

***(iii) Ensemble-based approaches: combining multiple PRSs***

The methods described above all share a common starting point: they take GWAS summary statistics as input and produce a single PRS. A more recent family of approaches takes a different perspective: they combine multiple PRSs that have already been constructed — either from different GWAS for the same trait, from different ancestries, or from genetically correlated traits — into a single, integrated score.

PRSmix and PRSmix+

PRSmix combines multiple PRSs developed for the *same* target trait, retrieved from the PGS Catalog. The combination is performed using Elastic Net regression in a development dataset, where each input PRS receives a *mixing weight* reflecting its independent contribution to predicting the trait. PRSmix+ extends this idea by additionally including PRSs from *other, genetically correlated traits (such as risk factors)*, thereby leveraging pleiotropy.

GPSMult and GPSMult+

GPSMult is a multi-ancestry, multi-trait ensemble approach that integrates genetic information across both ancestries and genetically related traits to predict a target trait of interest. It is constructed in two successive layers, each producing a usable score.
In the first layer, GPSMult combines ancestry-specific PRSs for the target trait — typically derived from GWAS conducted in different ancestry groups — into a single multi-ancestry, trait-specific score. The same procedure is applied independently to each genetically related risk factor or correlated trait, yielding one multi-ancestry score per trait. This first-layer score therefore captures ancestry-specific signals for the target trait alone, and can be used as a stand-alone multi-ancestry PRS.
In the second layer (GPSMult+), the multi-ancestry score for the target trait is then combined with the multi-ancestry scores of the related traits to produce a single, fully integrated PRS that captures both ancestry-specific signals and pleiotropic effects from related traits.
At each layer, the optimal set of component PRSs is selected using a stepwise procedure based on the Akaike Information Criterion (stepAIC), and their weights are estimated by logistic regression in a development dataset. The contrast between the first-layer and the two-layer outputs illustrates the respective contributions of multi-ancestry integration and pleiotropy: each step adds incremental gains in predictive accuracy, with the largest improvements typically observed in non-European populations, which are usually underserved by single-ancestry PRSs.

***(iv) Other approaches***

This last family groups together methods that do not fit neatly into the three previous categories. They share with the filtering and Bayesian approaches the fact that they operate at the level of individual SNPs — assigning one weight per variant — rather than combining pre-built PRSs as the ensemble methods do. What sets them apart is that each introduces a distinctive ingredient: a variable-selection penalty for Lassosum, automated model selection for MegaPRS, and explicit modelling of non-additive effects for GenoBoost

Lassosum

Lassosum belongs to a family of statistical techniques known as *penalized regression*, in which the model is forced to keep weights small unless the data strongly support a larger value. As with Bayesian methods, Lassosum does not start from the full GWAS but from a pre-defined panel of common, well-imputed variants (typically of the order of one million SNPs), intersected with those available in the GWAS summary statistics. Within this input panel, the model jointly estimates a weight for every variant from the GWAS summary statistics and an LD reference panel, while imposing a penalty — called the *LASSO* — that pulls weights toward zero. This penalty has a particularly useful property: it not only shrinks weak effects, but actually sets many of them *exactly* to zero. In practice, this means that the selection of SNPs is performed automatically by the model itself: variants with weak signals are dropped from the score, while those with strong signals are retained. The final weights are the non-zero penalized regression coefficients produced by the model — that is, the GWAS effect sizes re-estimated jointly under the LASSO penalty, not the original GWAS effect sizes. Two parameters control the procedure: λ (the strength of the penalty, which determines how many variants are kept) and *s* (which balances how much LD information is used). Both are tuned on the development dataset.

MegaPRS[16]

A practical difficulty with PRS construction is that no single method works best for all traits: some traits are driven by a small number of strong variants, while others involve many small contributions spread across the genome. The best statistical strategy depends on this underlying *genetic architecture*, which is usually unknown in advance. MegaPRS addresses this problem by trying several construction strategies on the same GWAS summary statistics — including penalized-regression and Bayesian approaches under various assumptions about how heritability is distributed across the genome — and automatically selecting the one that fits the data best. The selection of SNPs and the computation of their weights therefore depend on the winning strategy: if the best-fitting model is a Bayesian one, the final weights are the posterior mean effect sizes produced by that model on a pre-defined variant panel (every variant in the panel keeps a non-zero weight); if it is a penalized-regression model, the final weights are the non-zero penalized coefficients, and the SNP selection is performed automatically by the penalty. In either case, the user does not need to choose a method a priori: MegaPRS makes that choice based on the trait at hand, and returns the SNP weights produced by the winning strategy.

GenoBoost[17]

All the methods described so far assume that genetic variants act *additively* — meaning that carrying two copies of a risk allele increases risk exactly twice as much as carrying one copy. In reality, this is not always the case: for some variants, carrying just one copy may already confer almost the full risk (a *dominant* effect), while for others, risk increases only when both copies are present (a *recessive* effect). GenoBoost is designed to capture these non-additive patterns. Unlike all the methods described above, GenoBoost does not work from GWAS summary statistics but directly on individual-level genetic data, which is what allows it to estimate non-additive effects properly. The model is built step by step using a machine-learning technique called *gradient boosting*: at each iteration, GenoBoost scans all candidate variants, selects the single one that would most improve prediction given the variants already in the model, and adds it to the score. For each variant retained, GenoBoost does not assign a single weight, but three weights — one for each possible genotype (two copies of the major allele, one of each, two copies of the minor allele) — which are estimated from the individual-level data so as to maximize the model's fit to the trait. The data then determine whether the relationship between genotype and risk is additive, dominant, recessive, or something in between. The final SNPs included in the score are those selected across the iterations, and their weights are the three genotype-specific values estimated by the boosting procedure — not GWAS effect sizes. The number of iterations, the learning rate, and the level of regularization applied to homozygous minor alleles are all tuned on the development dataset. GenoBoost has been shown to outperform additive-only methods for autoimmune diseases such as rheumatoid arthritis and psoriasis, where non-additive effects play a substantial role — especially in the major histocompatibility complex (MHC) region of the genome.

**Detailed methodology**

***Step 1 — Does variability reflect additional predictive information or statistical uncertainty?***

First, we assessed whether the observed proportion of extreme opposite rankings across PRSs could be explained by chance alone. Specifically, we tested whether the level of disagreement between PRSs was greater than what would be expected if all PRSs were measuring the same underlying genetic risk, with differences arising only from random variation. To do this, we performed a Monte Carlo simulation in which we generated simulated PRS data that preserved the correlations observed between PRSs in the study population. Using these simulated data, we estimated the expected proportion of individuals who had at least one PRS in the top 5% and at least one PRS in the bottom 5% of the risk distribution, as well as analogous thresholds at the 10th and 90th percentiles. The expected proportions and their 95% simulation intervals were then compared with the observed values. If observed discordance exceeds simulated expectations, this would support the hypothesis that PRSs capture additional predictive information such as distinct biological pathways. Conversely, similar observed and expected values would support the hypothesis that variability arises from statistical uncertainty within a shared genetic signal. Additionally, we explored whether specific phenotypic characteristics were overrepresented among participants with discordant PRSs. If discordant PRSs reflect additional predictive information such as distinct biological pathways, individuals with discordant classifications would be expected to show specific phenotypic profiles.

Second, we quantified SNP-set similarity between PRSs using complementary metrics capturing different aspects of similarity. We calculated the Jaccard index, which measures overall overlap as the proportion of shared SNPs relative to the total number of SNPs across both PRSs. We also computed the SNP count ratio, which reflects how similar the PRSs are in size, independent of their overlap. Finally, we estimated the proportion of SNPs shared relative to each individual PRS, which captures whether one PRS is largely contained within another (i.e., nested relationships). We also assessed similarity in individual-level risk estimates using Pearson correlation coefficients. These metrics allowed us to distinguish between two scenarios: if PRS-specific SNPs capture additional predictive information, low SNP overlap should be associated with low correlation between PRSs. Conversely, similar individual-level behavior despite low overlap, particularly in the presence of nested structures, would suggest that variability is not primarily driven by distinct biological signals.

Third, we examined how much genetic information was shared across PRSs and how much was specific to each score, and whether these PRS-specific components provided additional predictive value beyond what was shared. To do this, we used linkage disequilibrium (LD) patterns estimated from genotype data in the CoLaus|PsyCoLaus cohort to identify groups of correlated genetic variants. Within each PRS, correlated variants were grouped into genomic regions, and a representative "index" SNP was selected to avoid counting the same genetic signal multiple times. For each pair of PRSs, variants were considered overlapping if they were located within 500 kb and moderately correlated (LD $r^2 > 0.2$), indicating that they likely represent the same underlying genetic signal; all others were considered unique. We then constructed separate scores capturing shared (overlapping) and PRS-specific (unique) components and jointly modeled them in logistic regression (ASCVD ~ Overlap_A + Overlap_B + Unique_A + Unique_B). This allowed us to test whether PRS-specific genetic information contributed to disease prediction beyond what was shared. To ensure reliable interpretation, we assessed multicollinearity using variance inflation factors (VIFs), as highly similar PRSs can produce correlated predictors. Statistical significance of unique components was evaluated using false discovery rate correction (FDR < 0.05) and interpreted in light of VIF values. We also quantified the relative contribution of shared versus unique components using Nagelkerke's $R^2$ and assessed correlations between components across individuals. If PRS-specific components contribute substantially to prediction beyond shared components, this would support the hypothesis that PRSs capture complementary risk information such as complementary biological pathways. Conversely, if prediction is mainly driven by shared components, this would indicate that most PRSs rely on a common underlying genetic signal.

***Step 2 —What drives intra-individual instability in PRS construction ?***

We first evaluated whether differences in SNP inclusion contributed to variability. If this is a major driver, PRSs with greater SNP overlap or more similar SNP-set sizes (i.e., whether two PRSs use a comparable number of genetic variants independently of their overlap; ratio of SNP counts ) should produce more similar individual risk estimates. We then assessed whether differences in SNP weighting contributed to variability by examining whether PRSs with identical or highly overlapping SNP sets could still produce different individual risk estimates. If such PRSs still show differences, this would indicate that variability arises from how SNP effects are weighted rather than which SNPs are included. We finally integrated features influencing SNP weighting—such as GWAS source, ancestry composition, and PRS construction method—into a multivariable pairwise model alongside SNP overlap and SNP set size. This approach allowed us to distinguish whether variability arises from differences in how genetic effects are estimated at the GWAS level (e.g., outcome definition or study population) or from how these effects are transformed and combined during PRS construction (e.g., Bayesian shrinkage, clumping, or ensemble methods).

***Step 3 — Does grouping PRSs by construction features reduce individual-level variability?***

Lastly, to determine whether specific aspects of PRS construction contribute to intra-individual variability, we grouped PRSs based on shared construction features, including GWAS source, ancestry composition, SNP set size, statistical method, and use of ensemble-based approaches. We then reassessed intra-individual variability within these groups to evaluate whether grouping PRSs with similar construction characteristics reduced variability and discordance at extreme percentiles. Based on these findings, we iteratively excluded PRSs whose construction features were associated with higher variability or frequent discordance and reassessed whether this stepwise refinement reduced overall variability. If variability is primarily driven by biological differences, grouping PRSs by construction features would be expected to have little effect, as distinct biological pathways would still be captured across scores. In contrast, if variability is driven by statistical and methodological factors, grouping PRSs with similar construction features should reduce variability and discordance. This step therefore also provides a practical framework to identify combinations of PRSs, and specific construction characteristics, that yield more consistent and clinically interpretable individual risk estimates.

### Supplementary Figures

**Supplementary Figure 1**. Flowchart

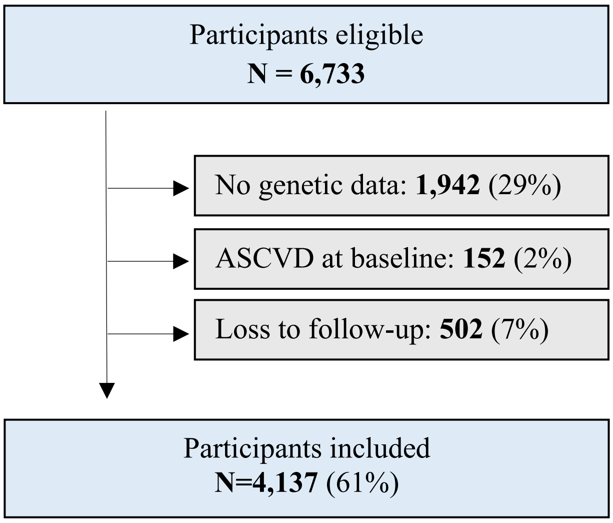

**Supplementary Figure 2.** (A) Brier scores (B) AUROC of all selected PRS

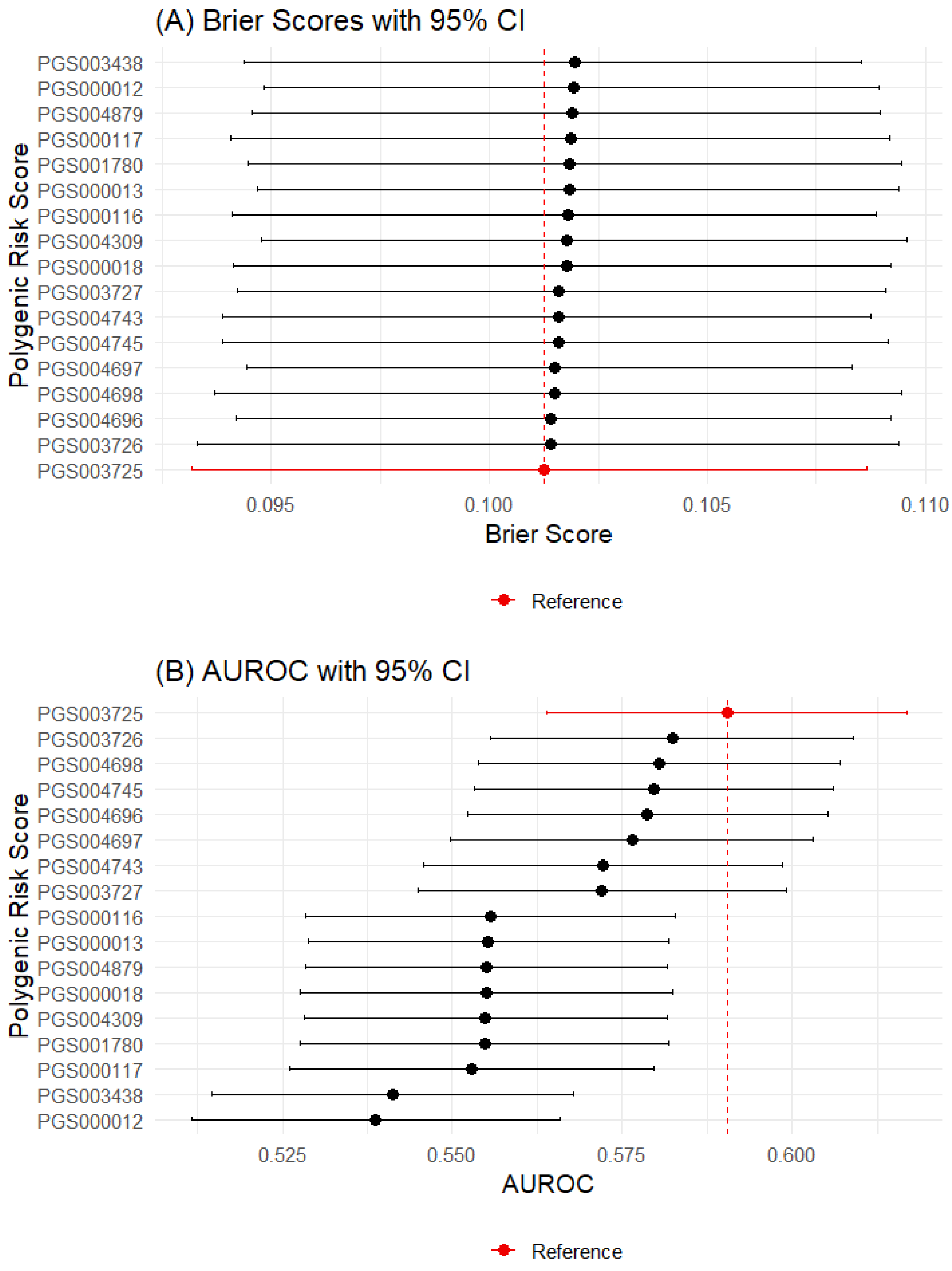

**Supplementary Figure 3. Nested structure of PRSs based on SNP overlap**

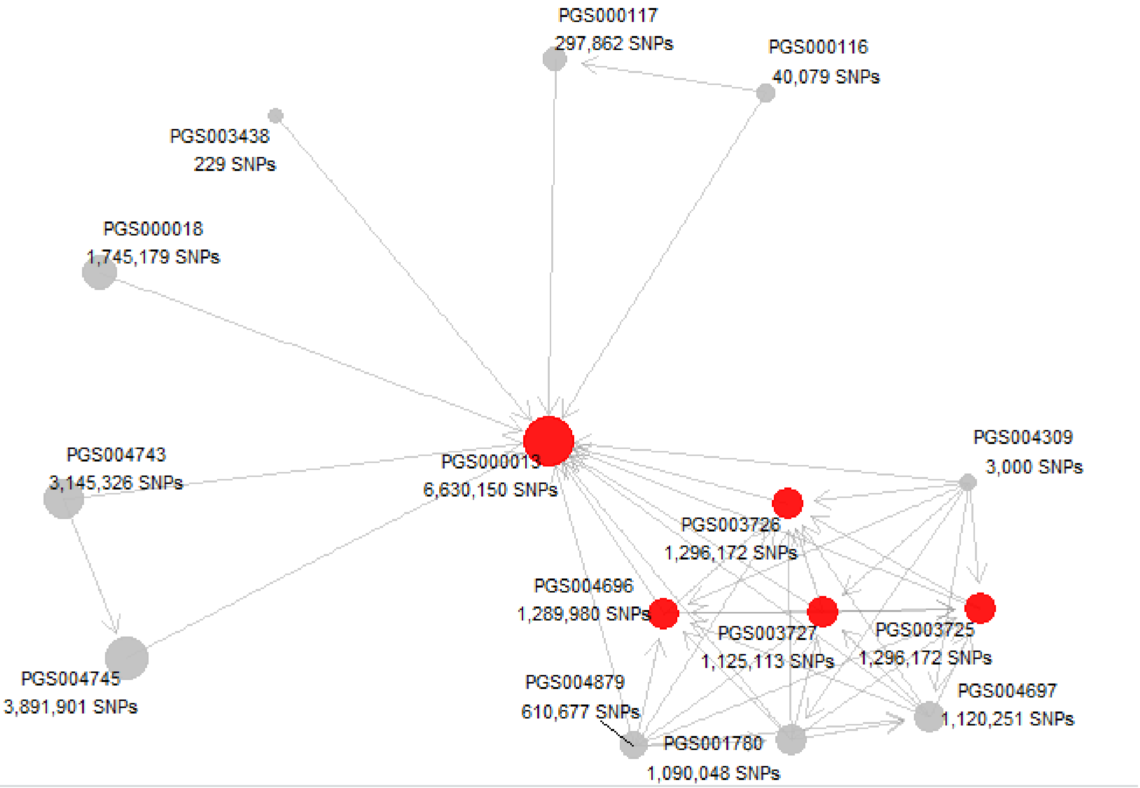

Each node represents a PRS, with node size proportional to the number of SNPs included. Directed edges indicate nesting relationships, pointing from the smaller (nested) PRS to the larger (container) PRS when ≥70% of the smaller PRS SNPs are shared across 42 out of 120 pairwise comparisons. Nodes highlighted in red denote PRSs with more than three incoming connections, indicating that they incorporate genetic information from multiple other scores. The network reveals a central hierarchical structure, with several PRSs (notably PGS003726, PGS003725, and PGS003727) acting as intermediate aggregators between smaller scores and the largest PRS (PGS000013).

**Supplementary Figure 4.** Decomposition of predictive variance into shared and PRS-specific genetic components for each PRS pair

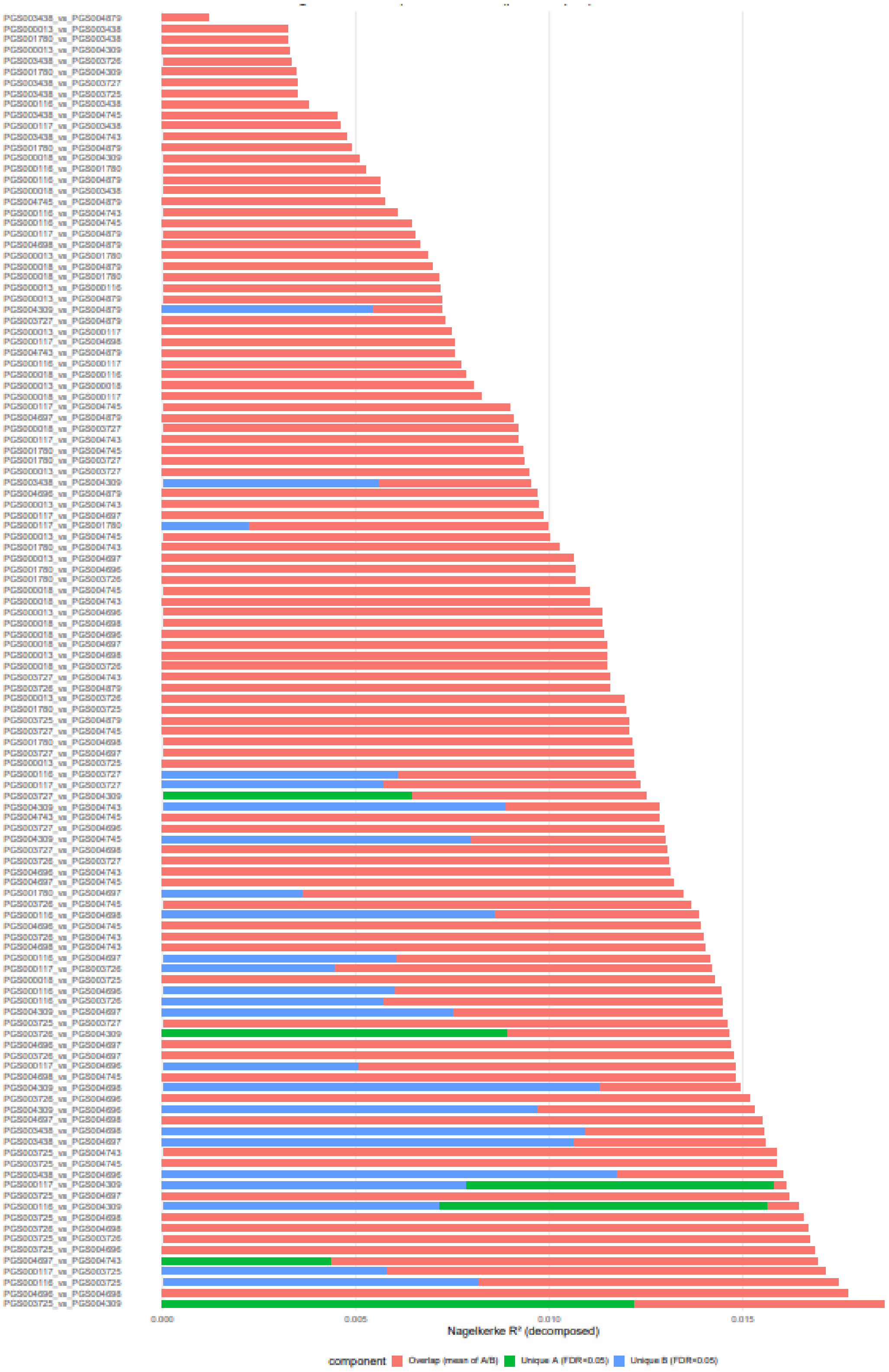

For each pair of ASCVD PRSs, stacked bars show the Nagelkerke $R^2$ attributable to the overlap component (shared genetic signal; mean of overlap terms) and to each FDR-significant PRS-specific unique component. PRS pairs are ordered by total decomposed $R^2$

**Supplementary Figure 5**. Redundancy diagnostics: overlap similarity vs. multicollinearity

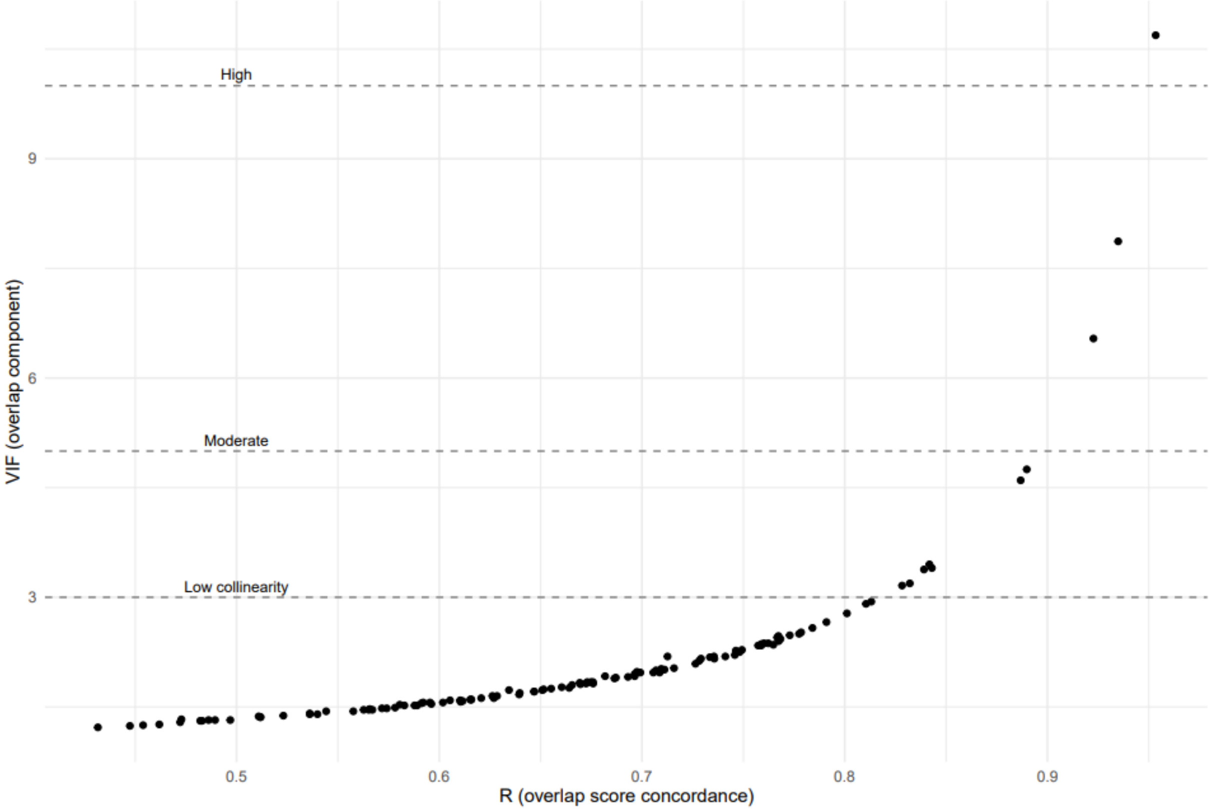

Variance inflation factors (VIFs) for the overlapping components of PRS pairs plotted against their overlap score concordance (R). This analysis evaluates the statistical stability of joint regression models by quantifying multicollinearity as a function of genetic similarity. As expected, higher concordance between PRSs is associated with increasing VIF values, reflecting greater statistical redundancy. Dashed horizontal lines indicate conventional thresholds for low, moderate, and high multicollinearity. Increasing VIF values mark a reduced ability to disentangle PRS-specific effects, thereby defining the range of genetic similarity over which score-specific interpretations remain statistically reliable. PRS pairs with high concordance ($r > 0.8$) showed substantially elevated VIFs, in some cases exceeding 5–10, reflecting considerable statistical redundancy. To ensure reliable component-level interpretation, statistical significance was not considered for the 5 PRS pairs with overlap concordance > 0.8 in the joint regression analyses.

**Supplementary Figure 6**. Frequency of PRS-specific independent predictive contributions across pairwise comparisons.

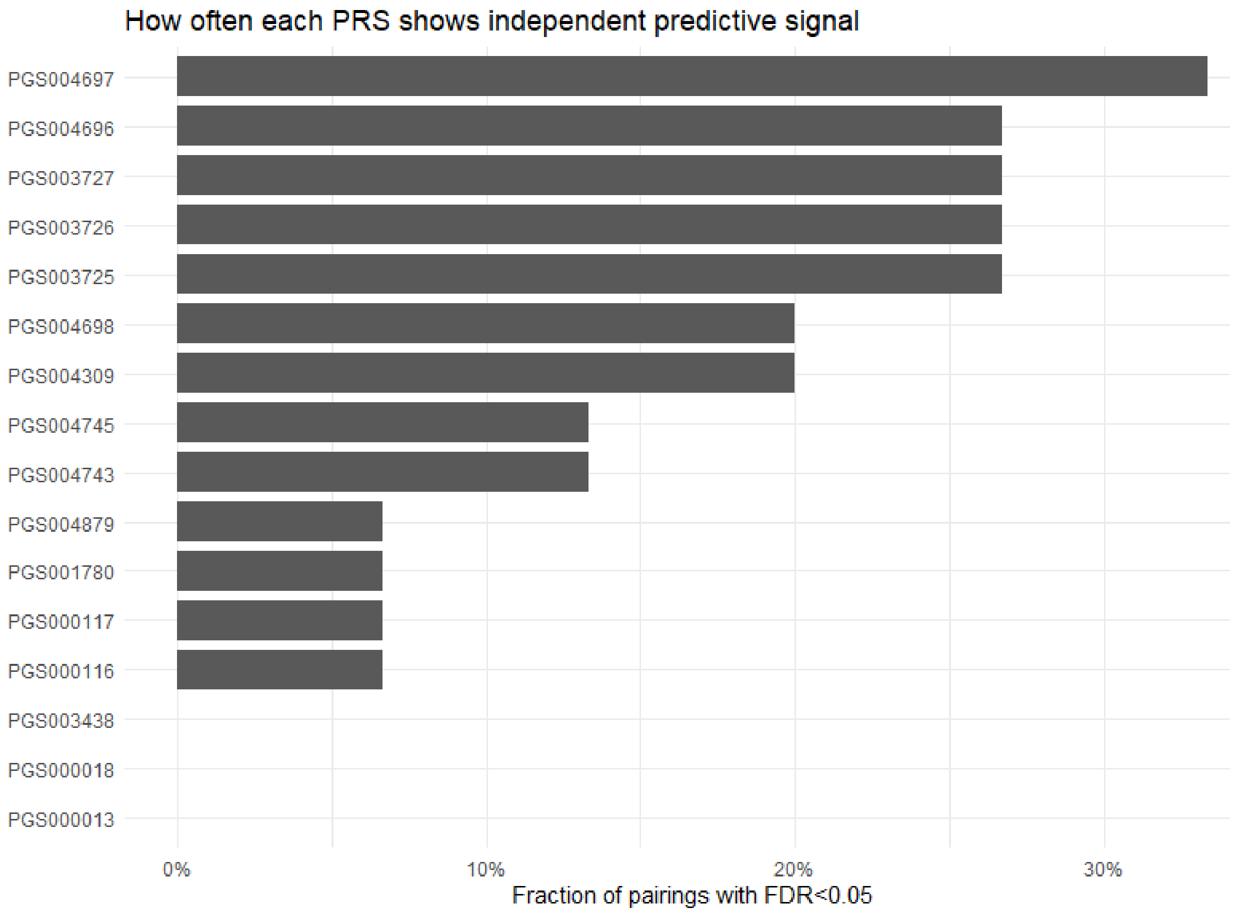

For each ASCVD polygenic risk score (PRS), the bar height represents the fraction of pairwise comparisons in which the PRS-specific (unique) component remained significantly associated with CVD status after false discovery rate correction (FDR < 0.05) in the joint regression model. Higher values indicate PRSs that more consistently provide independent predictive information beyond the shared genetic signal captured by other scores. PRSs are ordered by increasing fraction of significant pairings.

**Supplementary Figure 7.** Heatmap of weighted Cohen's Kappa across integrated scores (categorical)

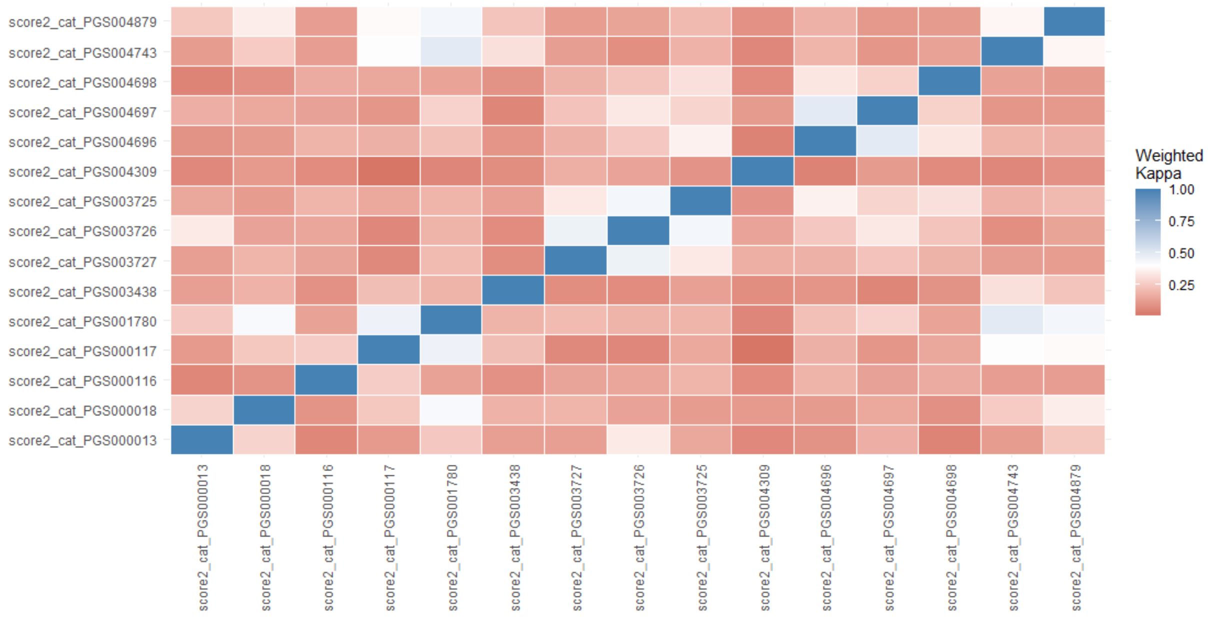

**Supplementary Figure 8.** Proximity to risk classification thresholds according to classification stability across 16 PRS-integrated SCORE2-OP models

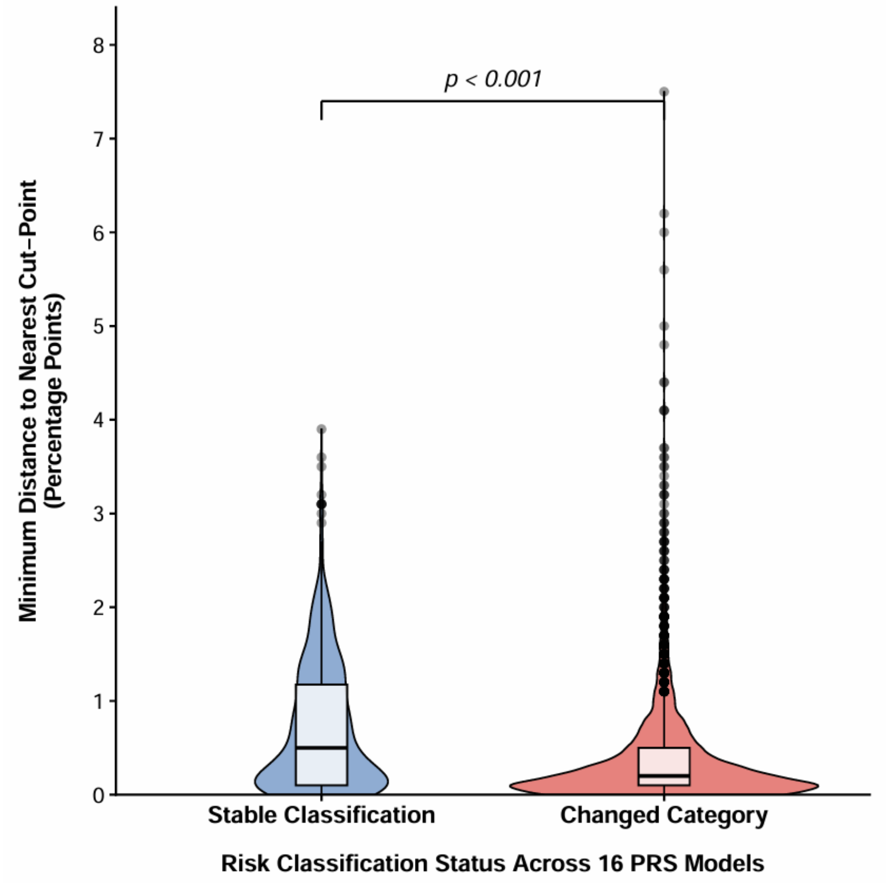
